# Socioeconomic Inequalities in Non-Adherence to Antihypertensive Medication in Peru

**DOI:** 10.1101/2024.06.12.24308773

**Authors:** Claudio Intimayta-Escalante, Lynn A. Quintana-García

**Affiliations:** Facultad de Medicina de San Fernando, Universidad Nacional Mayor de San Marcos, Lima, Peru; Facultad de Medicina Humana, Universidad Ricardo Palma, Lima, Peru

**Keywords:** Socioeconomic Health Disparities, Health Inequality Monitoring, Health Inequities, Hypertension, Treatment Adherence and Compliance, Antihypertensive Agents, Peru

## Abstract

**Introduction:** In Peru, adherence to antihypertensive treatment ranges from 55.5% to 46.6%. Adherence decreases under adverse socioeconomic conditions. Therefore, the aim of this research was to evaluate the socioeconomic inequalities in non-adherence to antihypertensive medication in Peru.

**Methods:** A cross-sectional study was conducted through the analysis of data from the Demographic and Family Health Survey carried out in Peru between 2013 and 2023. We addressed the socioeconomic conditions of hypertensive Peruvian adults, including sex, age group, educational level, area or place of residence, and health insurance. These conditions were evaluated as sources of inequality in non-adherence to antihypertensive medication at a general level using the Concentration Index (CI) or Erreygers’ Correction Index (ECI) among adults over 29 years of age.

**Results:** In the 15,624 hypertensive adults over 29 years old included in the study, the 86.63% had adherence to their antihypertensive medication. However, the inequality in non-adherence to medication was considerable (CI: -0.259; 95%CI: -0.385 to -0.132; p¡0.001). This inequality was greater among those aged 30 to 49 years (CI: -0.064; 95%CI: -0.121 to -0.008; p=0.01-0.2), those living in rural areas (CI: -0.484; 95%CI: -0.537 to -0.430; p¡0.001) or outside the capital (CI: -0.495; 95%CI: -0.569 to -0.422; p¡0.001), and those with no education (CI: -0.156; 95%CI: -0.201 to -0.112; p¡0.001) or only primary education (CI: -0.323; 95%CI: -0.386 to -0.260; p¡0.001). In contrast, hypertensive adults with a university education had a positive CI (CI: 0.257; 95%CI: 0.199 to 0.316; p¡0.001).

**Conclussion:** Hypertensive Peruvian adults residing in rural areas or outside the capital, and those with a low educational level, exhibited greater inequality in adherence to antihypertensive medication.

## INTRODUCTION

The World Health Organization (WHO) estimates that approximately 1.28 billion adults worldwide, aged between 30 and 79 years, suffer from hypertension (1). Additionally, out of every three adults with hypertension, two are from middle- and low-income countries. Only 42% of hypertensive adults receive medical treatment, while 21% have their blood pressure under control and 50% do not adhere to therapeutic guidelines (2), which can lead to the development of severe complications such as myocardial infarctions, cerebrovascular accidents, renal failure, and other health issues (3).

WHO provides a model with five dimensions that influence how well patients stick to their treatment: economic factors, healthcare system elements, treatment-related factors, disease-related factors, and personal factors (4,5). By 2025, it is projected that emerging countries in Latin America will have 1.56 billion cases of hypertension, coinciding with the World Health Assembly’s goal of reducing uncontrolled blood pressure by 25% (6).

In Peru, adherence to antihypertensive treatment affects quality of life, work productivity, and healthcare expenses (9). This lack of adherence highlights the necessity of introducing tailored health education initiatives and individualized support services to encourage proper treatment adherence (10). Additionally, factors such as lack of health knowledge, non-compliance with therapeutic guidelines, and medication costs contribute to this lack of adherence. Other influencing factors include aging, education, income, area of residence, healthcare coverage, and psychosocial conditions (11,12).

The hypertension management guidelines in Peru overlook patient adherence to treatment, worsening the task of guaranteeing efficient hypertension management and leading to less than optimal health results, revealing a notable flaw in the healthcare system (13,14). Neglecting sociodemographic variations in medication access and overlooking gaps in adherence awareness can severely impede treatment effectiveness, resulting in less than optimal health outcomes and difficulties in achieving successful hypertension management (15,16). Thus, the objective of this study is to assess socioeconomic inequalities in non-adherence to antihypertensive medication in Peru.

## METHODS

### Study Design

A cross-sectional study was conducted with an analysis of Demographic and Family Health Survey from Peru (or ENDES, in Spanish acronym). Peru is a country in Latin America with almost 32 million people (17). This study analyzed hypertensive adults who responded to the assessment on their antihypertensive medication adherence and had their blood pressure measured in the ENDES between 2013 and 2022.

### Variables Assessed

It was asked of hypertensive adults, “Did you take the medications as your doctor prescribed?”. This allowed the evaluation of medication adherence. Additionally, other variables were evaluated, including sex (male or female), age group (30-49, 50-64, and 65 or older), education level (without education, elementary, high school, or university), wealth quintile (first, second, third, fourth, and fifth quintile), area (rural or urban) or place of residence (living in or outside the capital), and health insurance affiliation (yes or no).

### Control of Blood Pressure

Among the hypertensive adults, systolic blood pressure (SBP) and diastolic blood pressure (DBP) were measured. In this way, considering the blood pressure ranges stipulated by clinical practice guidelines for hypertension management (18). We considered SBP values below 130mmHg and DBP values below 80mmHg to be healthy. Thus, we conducted an evaluation of medication adherence in hypertensive adults and the range of SBP/DBP they maintained. Additionally, we addressed the differences that exist according to the wealth quintile to evaluate inequality in medication adherence based on blood pressure.

### Statistical Analysis

The statistical analysis was conducted using R Studio version 4.2.2 (https://cran.r-project.org/), including the complex sample design inherent to ENDES. Categorical variables were described using frequencies and percentages, while numerical variables were presented as means with their respective 95% confidence intervals weighted by the design effect. Differences in medication intake among adults across different study variables were assessed using the Rao-Scott test. Thus, the association between sociodemographic characteristics and medication intake was assessed using Poisson regression models with robust variances to estimate both the crude Prevalence Ratio (PRc) and the adjusted Prevalence (aPR), accounting for all variables.

### Inequality Analysis

Inequality analysis was conducted using concentration index to assess the medication intake based on socioeconomic status, ranging from the fifth quintile to the first quintile. So, the concentration index (CI) was found by guessing the area above or below the curve. This way, conditions that generate clustering above the curve are linked to inequality caused by poverty (19). Additionally, Erreygers’ Concentration Index (ECI) was employed to allow for a more equitable estimation by considering the extremes of the distribution of the evaluated health condition (20). A map of Peru was created to show where hypertensive adults are located, their medication intake, and the ECI values in different regions.

### Ethical Aspects

Given that the ENDES data collection process involved participants’ informed consent, no ethical committee evaluation was necessary. Moreover, data obtained from the INEI platform are anonymized and securely stored (https://proyectos.inei.gob.pe/microdatos/).

## RESULTS

Of the 15,624 Peruvian adults aged over 29 years with arterial hypertension included in the study, 62.95% (95%CI: 61.49 to 64.39) were female, with a mean age of 64.44 years (95%CI: 63.97 to 64.85). Among the respondents, nearly half were aged 65 or older (51.83%, 95%CI: 50.25 to 53.39), had completed secondary education or higher (56.48%, 95%CI: 55.09 to 57.86), and belonged to the first two quintiles of wealth (52.61%, 95%CI: 51.01 to 54.22). Regarding place of residence, 43.38% (95%CI: 41.77 to 45.00) lived in the capital, while 84.11% (95%CI: 83.16 to 85.00) resided in urban areas. Concerning health insurance affiliation, 87.77% (95%CI: 86.55 to 88.90) were affiliated with some form of insurance. Meanwhile, 86.63% (95%CI: 85.46 to 87.72) adhered to the intake of antihypertensive medications as prescribed by their doctors.

Certain sociodemographic characteristics, such as age group, educational level, wealth quintile, and place of residence, mediated statistically significant differences (p<0.050) in the proportion of hypertensive adults taking medication for their condition (Table 1). Specifically, it was identified that hypertensive adults living in rural areas had a 5.61% higher prevalence of adherence to medication intake (PRa: 1.056; 95% CI: 1.007 to 1.108; p=0.025) compared to hypertensive adults in urban areas. Adults in the third, fourth, and fifth wealth quintiles, on the other hand, were less likely to take their antihypertensive medications (Table 2). This was because 4.76%, 6.29%, and 12.07% of those in those groups did not take their medications as prescribed. Also, compared to hypertensive adults aged 65 or older, those aged 50 to 64 years and 30 to 49 years had rates of 5.51% (PRa: 0.945; 95%CI: 0.914 to 0.976; p=0.001) and 6.85% (PRa: 0.931; 95%CI: 0.897 to 0.968; p=0.001) not taking their blood pressure medicines as prescribed.

**Table 1.** Sociodemographic characteristics of Peruvian adults with arterial hypertension according to treatment adherence.

| Variables | Hypertensive adults who did not take medications as prescribed (N=2252) |  | Adults with hypertension who took their medications as prescribed (N=13372) |  | P-value** |
| --- | --- | --- | --- | --- | --- |
|  | n | %* (95%CI) | n | %* (95%CI) |  |
| **Sex?** |  |  |  |  |  |
| Male | 796 | 12.08 (10.58 a 13.76) | 4,716 | 87.92 (86.24 a 89.42) | 0.070 |
| Female | 1,456 | 14.13 (12.67 a 15.72) | 8,656 | 85.87 (84.28 a 87.33) |  |
| **Age Group?** |  |  |  |  |  |
| 30 to 49 years | 651 | 17.31 (15.24 a 19.60) | 2,681 | 82.69 (80.40 a 84.76) | <0.001 |
| 50 to 64 years | 782 | 16.06 (13.79 a 18.63) | 4,255 | 83.94 (81.37 a 86.21) |  |
| 65 or more years | 819 | 10.49 (9.16 a 11.98) | 6,436 | 89.51 (88.02 a 90.84) |  |
| **Educational Level?** |  |  |  |  |  |
| Without Education | 291 | 13.99 (11.60 a 16.78) | 1,588 | 86.01 (83.22 a 88.40) | <0.001 |
| Elementary | 808 | 12.86 (11.48 a 14.37) | 5,184 | 87.14 (85.63 a 88.52) |  |
| High School | 727 | 16.04 (13.90 a 18.43) | 3,632 | 83.96 (81.57 a 86.10) |  |
| University | 425 | 10.54 (9.04 a 12.26) | 2,964 | 89.46 (87.74 a 90.96) |  |
| **Wealth Index?** |  |  |  |  |  |
| Q5 (lower) | 631 | 17.31 (15.04 a 19.85) | 3,039 | 82.69 (80.15 a 84.96) | <0.001 |
| Q4 | 511 | 15.36 (13.52 a 17.40) | 2,812 | 84.64 (82.60 a 86.48) |  |
| Q3 | 458 | 14.85 (12.59 a 17.44) | 2,481 | 85.15 (82.56 a 87.41) |  |
| Q2 | 371 | 13.72 (10.91 a 17.13) | 2,423 | 86.28 (82.87 a 89.09) |  |
| Q1 (higher) | 281 | 9.44 (7.58 a 11.70) | 2,617 | 90.56 (88.30 a 92.42) |  |
| **Area of Residence?** |  |  |  |  |  |
| Urbano | 1,906 | 14.54 (13.04 a 16.18) | 10,817 | 85.46 (83.82 a 86.96) | 0.180 |
| Rural | 346 | 13.15 (11.89 a 14.51) | 2,555 | 86.85 (85.49 a 88.11) |  |
| **Place of Residence?** |  |  |  |  |  |
| Other Regions | 712 | 14.58 (13.31 a 15.96) | 3,763 | 85.42 (84.04 a 86.69) | 0.025 |
| Capital | 1,540 | 11.78 (9.97 a 13.87) | 9,609 | 88.22 (86.13 a 90.03) |  |
| **Health Insurance?** |  |  |  |  |  |
| No | 313 | 15.88 (13.09 a 19.14) | 1,387 | 84.12 (80.86 a 86.91) | 0.055 |
| Yes | 1,782 | 12.87 (11.65 a 14.18) | 11,189 | 87.13 (85.82 a 88.35) |  |

\* Estimated percentage considering the weighting factor.
\*\*P-value estimated with the Rao-Scott test.

**Table 2.** Regression models to estimate the prevalence of treatment adherence according to sociodemographic and health characteristics among hypertensive adults.

| Variables | Crude Model |  |  | Adjusted Model |  |  |
| --- | --- | --- | --- | --- | --- | --- |
|  | cPR | 95%CI | P-value* | aPR | 95%CI | P-value* |
| **Sex?** |  |  |  |  |  |  |
| Male |  | **REF** |  |  | **REF** |  |
| Female | 1.17 | 0.987 a 1.386 | 0.07 | 0.985 | 0.96 1.011 | 0.258 |
| **Age Group?** |  |  |  |  |  |  |
| 65 or more years |  | **REF** |  |  | **REF** |  |
| 50 to 64 years | 1.532 | 1.252 a 1.874 | <0.001 | 0.945 | 0.914 0.976 | 0.001 |
| 30 to 49 years | 1.65 | 1.374 a 1.983 | <0.001 | 0.931 | 0.897 0.968 | <0.001 |
| **Educational Level?** |  |  |  |  |  |  |
| University |  | **REF** |  |  | **REF** |  |
| High School | 1.521 | 1.256 a 1.842 | <0.001 | 0.956 | 0.927 0.986 | 0.004 |
| Elementary | 1.219 | 1.016 a 1.463 | 0.033 | 0.99 | 0.962 1.018 | 0.479 |
| Without Education | 1.327 | 1.058 a 1.665 | 0.014 | 0.98 | 0.939 1.022 | 0.338 |
| **Wealth Index?** |  |  |  |  |  |  |
| Q5 (lower) |  | **REF** |  |  | **REF** |  |
| Q4 | 1.454 | 1.061 a 1.993 | 0.020 | 0.966 | 0.929 1.005 | 0.083 |
| Q3 | 1.573 | 1.201 a 2.062 | 0.001 | 0.952 | 0.918 0.989 | 0.010 |
| Q2 | 1.628 | 1.267 a 2.091 | <0.001 | 0.937 | 0.901 0.974 | 0.001 |
| Q1 (higher) | 1.834 | 1.417 a 2.374 | <0.001 | 0.879 | 0.822 0.941 | <0.001 |
| **Area of Residence?** |  |  |  |  |  |  |
| Urbano |  | **REF** |  |  | **REF** |  |
| Rural | 1.106 | 0.954 a 1.282 | 0.180 | 1.056 | 1.007 1.108 | 0.025 |
| **Place of Residence?** |  |  |  |  |  |  |
| Other Regions |  | **REF** |  |  | **REF** |  |
| Capital | 0.808 | 0.669 a 0.975 | 0.027 | 1.006 | 0.977 1.037 | 0.682 |
| **Health Insurance?** |  |  |  |  |  |  |
| No |  | **REF** |  |  | **REF** |  |
| Yes | 0.81 | 0.655 a 1.002 | 0.053 | 1.022 | 0.982 1.063 | 0.288 |

**cPR:** crude Prevalence Ratio, **aPR:** Prevalence Ratio adjusted for the other variables, **95%CI:** Confidence interval at 95%, **REF:** Category used as reference for estimates.
\*P-value estimated by Poisson regression model with robust variance and weighting factor.

A big difference was seen between adults with high blood pressure who didn’t take their medicine as prescribed (CI: -0.259; 95%CI: -0.385 to -0.132; p<0.001) and those who did take their antihypertensive medicine as prescribed (CI: 0.038; 95%CI: 0.012 to 0.063; p=0.004) (Figure 1A). Also, among hypertensive adults who were taking their medications, there was more inequality among those with only elementary education (CI: -0.287; 95%CI: -0.373 to -0.200; p<0.001), living in rural areas (CI: -0.853; 95%CI: -1.031 to -0.675; p<0.001), or not living in the capital (CI: -0.441; 95%CI: -0.631 to -0.251; p<0.001), compared to those with a university education (CI: 0.257; 95%CI: 0.199 to 0.316; p<0.001). Also, among adults with high blood pressure who didn’t take their medicine as prescribed, those with only primary education (CI: -0.279; 95% CI: -0.538 to -0.019; p=0.036) and living in rural areas (CI: -0.976; 95%CI: -1.426 to -0.527; p<0.001) or outside the capital (CI: -0.402; 95%CI: -0.596 to -0.207; p<0.001) were more unequal (Figure 2A) than those with a university education (CI: 0.508; 95%CI: 0.257 to 0.758; p<0.001).

**Figure 1.**
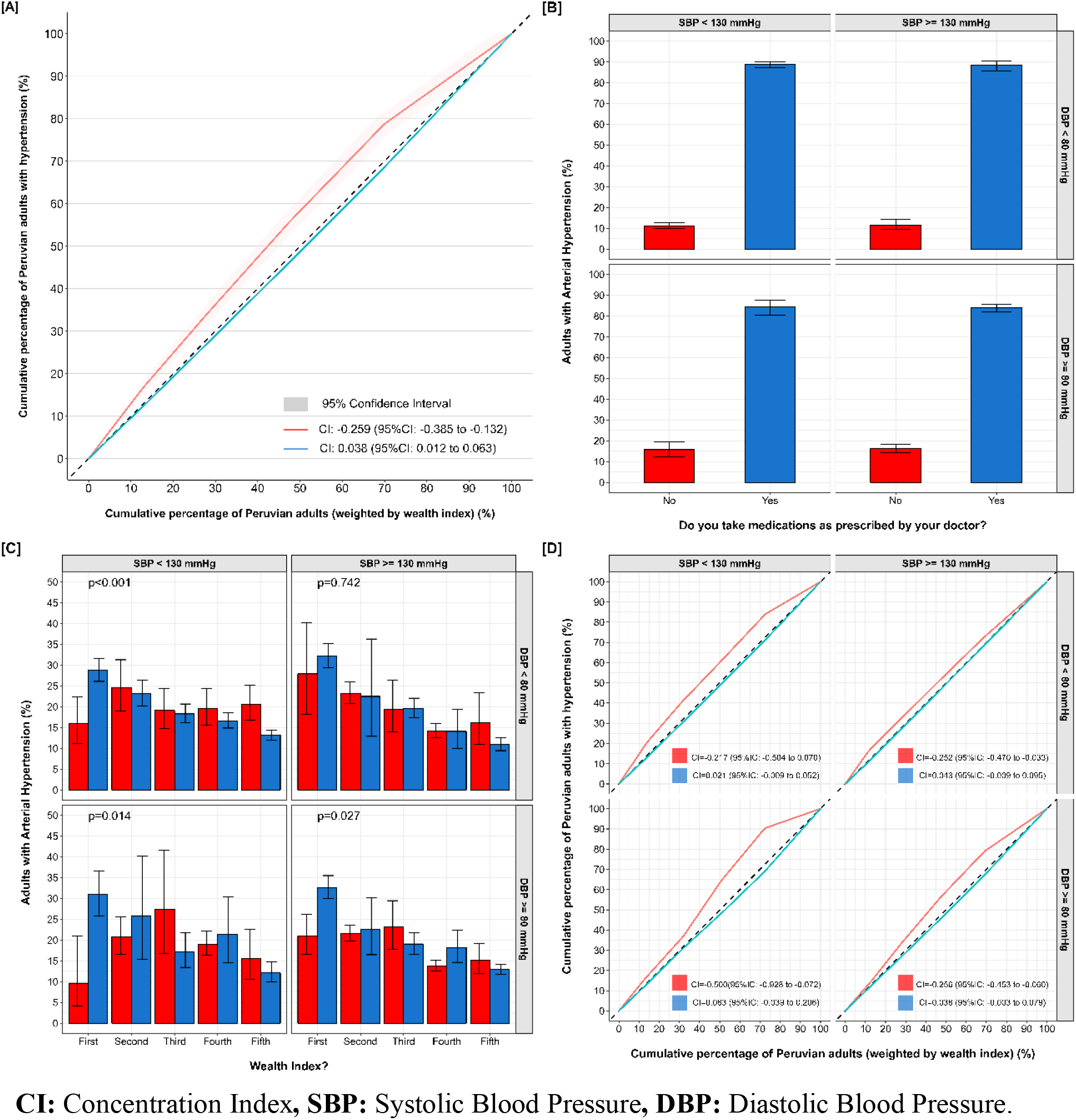
Socioeconomic inequality in adherence to antihypertensive treatment according to blood pressure range in Peruvian adults

**Figure 2.**
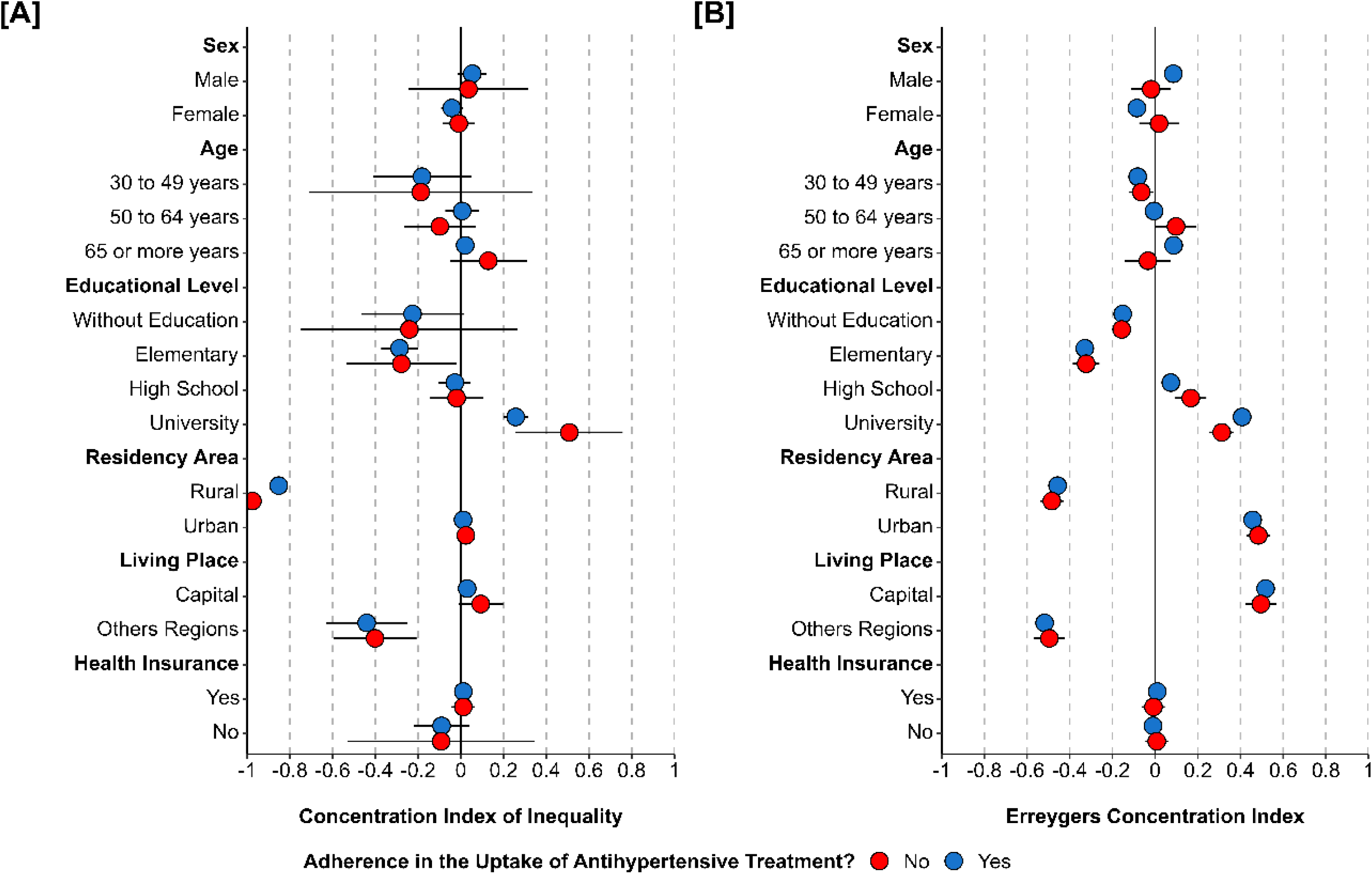
Socioeconomic inequality in adherence to antihypertensive treatment in Peruvian adults

In evaluating inequality with the Erreygers Concentration Index (ECI), it was identified that among hypertensive adults with adherence to medication intake, women (ECI: -0.085; 95% CI: -0.118 to -0.052; p<0.001), those aged 30 to 49 years (ECI: -0.082; 95%CI: -0.103 to -0.061; p<0.001) and 65 or older (ECI: 0.087; 95% CI: 0.052 to 0.122; p<0.001), without education (ECI: -0.152; 95%CI: -0.171 to -0.133; p<0.001) or only elementary education (ECI: -0.329; 95%CI: -0.354 to -0.304; p<0.001), living in rural areas (ECI: -0.457; 95%CI: -0.483 to -0.431; p<0.001) or regions outside the capital (ECI: -0.518; 95%CI: -0.549 to -0.486: p<0.001) exhibited greater inequality. Also, among adults with high blood pressure who didn’t take their medicine as prescribed, those aged 30 to 49 (ECI: -0.064; 95%CI: -0.121 to -0.008; p=0.012), without education (ECI: -0.156; 95%CI: -0.201 to -0.112; p<0.001) or elementary education (ECI: -0.323; 95%CI: -0.386 to -0.260; p<0.001), living in rural areas (ECI: -0.484; 95%CI: -0.537 to -0.430; p<0.001) or outside the capital (ECI: -0.495; 95%CI: -0.569 to -0.422; p<0.001) showed more inequality (Figure 2B).

In the assessment of blood pressure among hypertensive Peruvian adults, an average systolic blood pressure (SBP) of 143.34 mmHg (95%CI: 142.64 to 144.07) and an average diastolic blood pressure (DBP) of 77.81 mmHg (95% CI: 77.40 to 78.23) were identified. Additionally, it was found that among hypertensive Peruvian adults, only 26.40% (95%CI: 25.09 to 27.74) fell within the optimal range for SBP<130 mmHg and DBP<80 mmHg. Within this group, 88.81% (95%CI: 87.24 to 90.21) adhered to medication intake. Conversely, 36.92% (95%CI: 35.44 to 38.43) were outside the optimal blood pressure range (SBP>129mmHg/DBP>79mmHg). It was found that within this group, only 83.86% (95%CI: 81.85 to 85.68) were adherent to medication intake (Figure 1B). Also, it was found that in both cases, the wealth quintile made a big difference in the number of adults with high blood pressure who took their medicine as prescribed (p<0.050) (Figure 1C). When inequality was looked at, it was found that adults with high blood pressure who didn’t take their medicine as prescribed and whose blood pressure readings were outside the ideal range had more inequality. This got worse as their readings moved away from the range of DBP>79mmHg and toward the ideal range of SBP<130mmHg (Figure 1D).

Across the 25 regions of Peru, although the frequency of hypertensive adults over 29 years old was less than 20% (Figure 3A), adherence within this group was over 70% (Figure 3B). However, regions in the Peruvian jungle and highlands exhibited greater inequality in adherence to antihypertensive medication intake (Figure 3C). Additionally, during the evaluated period, a slight decrease in the proportion of hypertensive Peruvian adults over 29 years old was identified from 2013 (17.30%, 95%CI: 15.28 to 19.53) to 2023 (14.93%, 95%CI: 14.06 to 15.85). Although the proportion of those adhering to medication remained consistent above 80%, ranging from 83.40% (95%CI: 72.74 to 90.43) to 89.50% (95%CI: 87.26 to 91.46).

**Figure 3.**
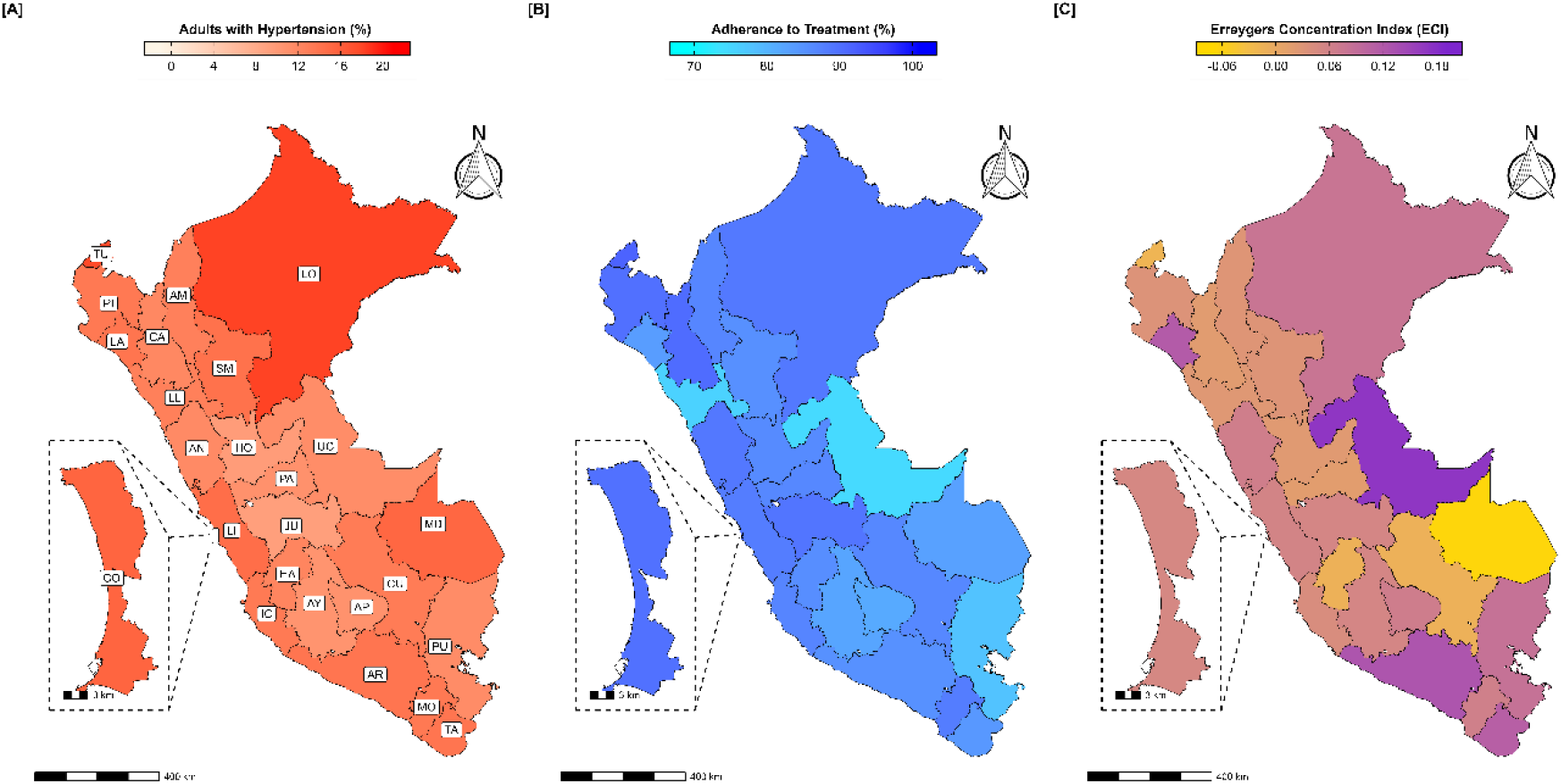
Distribution of adherence to antihypertensive treatment in Peruvian adults

Furthermore, a growing gap was evidenced in the number of hypertensive adults with non-adherence to medication intake in recent years (Figure 4A). On the other hand, when looking at the annual change in inequality among adults with high blood pressure who are taking their medications, it was found that the inequality indices got worse over time (Figure 4B).

**Figure 4.**
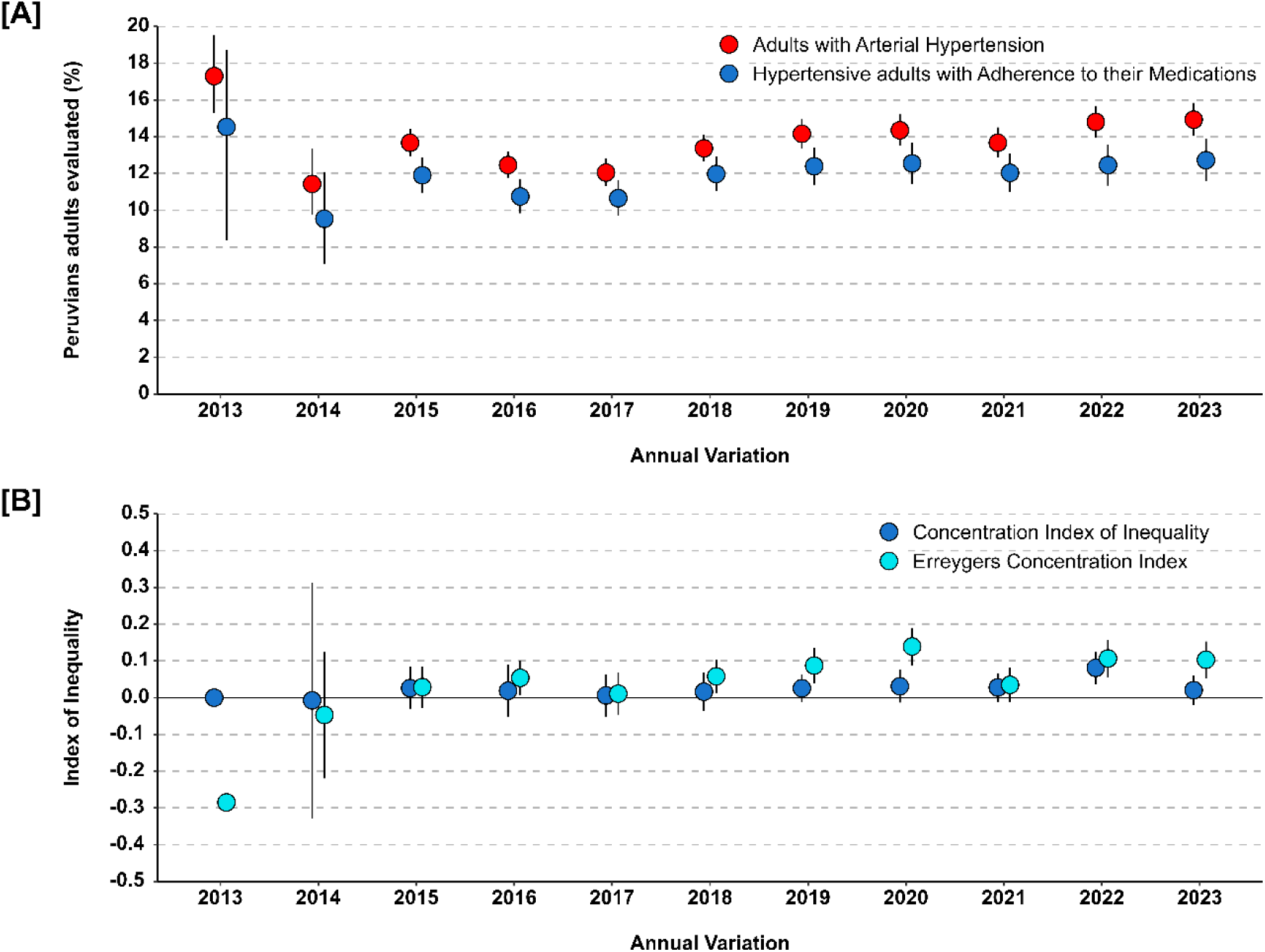
Annual variation of inequality in adherence to antihypertensive treatment in Peru

## DISCUSSION

This research assessed sociodemographic inequalities in medication intake among hypertensive adults in Peru. It was found that hypertensive adults in rural areas face greater disparities in medication intake. This is because there are few healthcare centers in rural areas, and access to antihypertensive medications is limited. In Peru, only 36% of public health centers have these medications available (21). This worsens the situation for the 23.1% of Peruvians without health insurance (22). This situation is similar to other Latin American nations, such as Brazil, where 10% of individuals with hypertension and diabetes do not have access to necessary medications for their conditions (23,24).

In this context, community health initiatives such as home monitoring, lifestyle counseling, and follow-up calls have shown efficacy in improving treatment adherence and identifying access issues to medications (25,26). However, in Peru, telemedicine faces challenges in incorporating digital interventions due to the lack of internet access, especially in rural areas like the jungle and highlands (27,28). As a result, interventions derived from this service for populations with chronic diseases are confined to urban areas (29). On the other hand, educational interventions targeting healthcare professionals and patients, combined with appointment reminders, have shown success (30,31).

Hypertensive adults with higher levels of education experience less inequality in medication intake, highlighting the vital role of education in comprehending cardio-metabolic diseases. This emphasizes the importance of education in understanding cardio-metabolic diseases (32). Individuals with a university education are better equipped to understand the significance of maintaining regulated “blood pressure levels” (33). Conversely, individuals with lower educational levels, particularly in populations with other languages and in rural areas, face greater challenges in following therapeutic guidelines (34,35). This is evident in Peruvian hospitals, where up to 35% of hypertensive adults struggle to understand health information, leading to only 15% following their treatment (36).

Among younger hypertensive Peruvian adults, lower inequality in medication adherence was observed, possibly due to the lower prevalence of formal employment among this age group (9,37). This may be because only one in four young adults in Peru has formal employment, which makes it difficult to integrate arterial hypertension treatment (9,38,39). Non-adherence to hypertension medication can lead to complications and an increased need for medication to control blood pressure (40,41). As the hypertensive population ages, they typically acquire greater knowledge about their condition and show increased treatment adherence (33,42). Integrating newly diagnosed hypertensive adults into support groups with those who have lived with the disease for longer periods could help reduce disparities in adherence to antihypertensive medications (43).

The lower occurrence of hypertension in women may clarify the variance in adherence to antihypertensive medication between men and women (44). However, it is important to consider that underreporting of hypertension in women may distort the assessment of inconsistent adherence to antihypertensive treatment. However, evidence indicates that women are at higher risk of developing hypertensive disorders in specific contexts, such as pregnancy and menopause (45-47). In the Peruvian context, there is no clinical-therapeutic approach tailored to these risks, which could lead to inadequate therapeutic regimens with insufficient antihypertensive medication, exacerbating adverse socioeconomic conditions (48-50).

Non-adherence in hypertensive patients with normal blood pressure values may stem from forgetfulness, a lack of motivation to maintain normal values, the presence of other health conditions, and perceiving the disease as not severe (51,52). Psychosocial intervention models help emphasize the benefits of treatment and provide skills for adherence, such as keeping medication in a visible place or taking it with breakfast (53). Additionally, setting shared goals between the physician and the patient has been shown to improve adherence (54).

The focus of this research is on examining differences in adherence to antihypertensive medication. However, the study has limitations due to the nature of the Demographic and Family Health Survey in Peru, which addresses hypertension or elevated blood pressure without delving into the context of diagnosis or assessment. Furthermore, the survey generally inquires whether adults take the medications prescribed by their doctors for hypertension or elevated blood pressure without evaluating the reasons, difficulties, and complications that might prevent them from adhering to their treatment properly. Similarly, it does not explore the knowledge of hypertensive adults about their disease or the importance of taking antihypertensive medication.

In conclusion, among hypertensive Peruvian adults, there are socioeconomic inequalities in adherence to medication for their condition. These inequalities are more pronounced among adults living in rural areas or outside the capital, those with lower educational levels, or those belonging to a younger age group. This disparity has decreased in specific regions of Peru in recent years. Identifying these sources of inequality helps pinpoint areas for enhancing antihypertensive treatment strategies in Peru, aiming to provide more equitable care for adults with hypertension.

## Data Availability

The data was obtained from the National Institute of Statistics and Informatics (or INEI, in Spanish) platform: https://proyectos.inei.gob.pe/microdatos/

## Acknowledgements

We thank the National Institute of Neoplastic Diseases in Peru for the development of the Demographic and Family Health Survey.

